# Severe Acute Respiratory Syndrome Coronavirus 2 (SARS-CoV-2) Infection During Pregnancy In China: A Retrospective Cohort Study

**DOI:** 10.1101/2020.04.07.20053744

**Authors:** Ming-Zhu Yin, Li-juan Zhang, Guang-Tong Deng, Chao-Fei Han, Min-Xue Shen, Hong-Yin Sun, Fu-Rong Zeng, Wei Zhang, Lan Chen, Qing-Qing Luo, Du-Juan Yao, Min Wu, Shi-Huan Yu, Hui Chen, David Baud, Xiang Chen

## Abstract

**Background:** Severe acute respiratory syndrome coronavirus 2 (SARS-CoV-2) has been identified as the cause of the ongoing worldwide epidemic of Coronavirus Disease 2019 (COVID-19) in China and worldwide. However, there were few studies about the effects of SARS-CoV-2 infection on pregnant women.

**Methods:** In this retrospective cohort study, we enrolled 31 pregnant women and 35 non-pregnant women from Jan 28 to Feb 28, 2020 to evaluate the effects of SARS-CoV-2 infection during pregnancy. Inflammatory indices were used to assess the severity of COVID-19. Evidence of vertical transmission was determined by laboratory confirmation of SARS-CoV-2 in amniotic fluid, placenta, neonatal throat and anal swab and breastmilk samples.

**Findings:** Compared with non-pregnant women, pregnant women had a significantly lower proportion of fever (54·8% vs. 87·5%, *p*= 0.006), a shorter average interval from onset to hospitalization (7·80 ±7·0d vs. 13·2 ± 8·2d, *p*= 0.005), and a higher proportion of severe or critical COVID-19 (32·3% vs. 11·4%, *p*=0.039). Neutrophil-to-lymphocyte ratio (NLR) and systematic immune-inflammation-based prognostic index (SII) were significantly higher on admission in severe/critical pneumonia group than moderate pneumonia group. We could not detect the presence of SARS-CoV-2 by RT-PCR in amniotic fluid, placenta, neonatal throat and anal swab and breastmilk samples.

**Interpretation:** The clinical symptoms of COVID-19 in pregnant women were insidious and atypical, compared with those in non-pregnant patients. SII and NLR could be a useful marker to evaluate the severity of COVID-19. There was no evidence of vertical transmission during pregnancy with SARS-CoV-2 infection.

**Funding:** National Natural Science Foundation of China and Research Funds for the Central Universities.

**Research in context:** *Evidence before this study:* We searched PubMed, Embase and Web of science for articles published up to March 1st, 2020, using the keywords (“novel coronavirus” OR “2019 novel coronavirus” OR “2019-nCoV” OR COVID-19 OR SARS-CoV-2) AND (pregnancy OR “maternal infection” OR “fetal infection”) AND “Cohort studies”. We identified no published cohort studies on pregnant women with the 2019 novel coronavirus disease (COVID-19) infection.

*Added value of this study:* For this retrospective cohort study, we reviewed clinical records, laboratory findings, and chest CT scans from 31 pregnant women and 35 non-pregnant women from Jan 28 to Feb 28, 2020 to evaluate the effects of SARS-CoV-2 infection during pregnancy. Inflammatory indices were used to assess the severity of COVID-19. Evidence of vertical transmission was determined by laboratory confirmation of SARS-CoV-2 in amniotic fluid, placenta, neonatal throat and anal swab and breastmilk samples. Compared with non-pregnant women, pregnant women had a significantly lower proportion of fever (54·8% vs. 87·5%, p= 0.006), a shorter average interval from onset to hospitalization (7·80 ±7·0d vs. 13·2 ± 8·2d, p= 0.005), and a higher proportion of severe or critical COVID-19 (32·3% vs. 11·4%, p=0.039). Neutrophil-to-lymphocyte ratio (NLR) and systematic immune-inflammation-based prognostic index (SII) were significantly higher on admission in severe/critical pneumonia group than moderate pneumonia group. Amniotic fluid, placenta, neonatal throat and anal swab and breastmilk samples were tested for SARS-CoV-2 by RT-PCR and all results were negative.

*Implications of all the available evidence:* The clinical symptoms of COVID-19 in pregnant women were insidious and atypical, compared with those in non-pregnant patients. SII and NLR could be a useful marker to evaluate the severity of COVID-19. There was no evidence of vertical transmission during pregnancy with SARS-CoV-2 infection.

## Introduction

Severe acute respiratory syndrome coronavirus 2 (SARS-CoV-2) has been identified as the cause of the ongoing worldwide epidemic of Coronavirus Disease 2019 (COVID-19) first in China and then worldwide.^1-5^ As of March 9, 2020, a total of 113, 702 cases had been reported, including 4, 012 deaths globally.^6^ Phylogenetic analysis revealed that SARS-CoV-2 was a novel single-stranded ribonucleic acid (RNA) betacoronavirus which resembled severe acute syndrome coronavirus (SARS-CoV)^5^. However, SARS-CoV-2 is far more severe and infectious than SARS-CoV, and has become a global health emergency.^4,7-9^

Although numerous studies have illuminated the clinical characteristics and outcomes of general population with COVID-19,^2,8^ little has been reported about the effects of SARS-CoV-2 infection on pregnant women.^10,11^ To our knowledge, a study by Huang and colleagues reported that the clinical characteristics of nine pregnant women with COVID-19 resembled those in the general population.^12^ Another study including ten pregnant patients demonstrated that perinatal SARS-CoV-2 infection had adverse effects on newborns such as fetal distress and even death.^13^ Moreover, a case described that a pregnant woman with COVID-19 delivered a healthy neonate with no evidence of SARS-CoV-2 infection during her 30 weeks pregnancy.^14^ However, these series of cases had limitations for their small size and lack of control group. Therefore, there are still debates on whether pregnant women have a different disease course and outcomes, considering the physiological changes in cell-mediated immunity and pulmonary function during pregnancy.^15^

In this study, we conducted retrospective cohort study to compare the effects of SARS-CoV-2 infection on pregnant women and non-pregnant women. We hope these findings will facilitate efforts, both in China and globally, to make and manage public health planning for pregnant women with COVID-19.

## Materials and methods

### Study design and participants

For this retrospective cohort study, we included two cohorts of female inpatients (20-40 years old, female) from Jan 28 to Feb 28, 2020, at Wuhan Union and Tongji hospitals of Huazhong University of Science and Technology. All female patients were diagnosed as COVID-19 based on the New Coronavirus Pneumonia Prevention and Control Program (6th edition) published by the National Health Commission of China.^16^ A total of 68 patients were initially enrolled, and 2 patients were excluded from our study because they had tumors with COVID-19 (one had cervical cancer and the other had lymphoma in non-pregnant group). Finally, we recruited 66 hospitalized patients with COVID-19 including 31 pregnant women and 35 non-pregnant women. All the patients were PCR-confirmed cases in our study. The clinical outcomes were monitored up to March 8, 2020, the last day of follow-up. Ethical approval for the study was obtained from the Ethics Committee of Wuhan Union and Tongji hospitals of Huazhong University of Science and Technology, and written informed consent was waived by the Ethics Commission of the designated hospital for the emergency of COVID-19.

### Data collection

We obtained demographic, clinical, laboratory, and radiological data from electronic medical records with data collection form. Two study investigators (CH, LZ) independently collected and validated the data. Maternal throat swab samples from the upper respiratory tract were collected at admission in viral-transport medium. Amniotic fluid samples, placenta samples, neonatal throat and anal swab samples were collected immediately after delivery in the operating room. Breastmilk samples were also collected for SARS-CoV-2 testing to evaluate the evidence of vertical transmission. The presence of SARS-CoV-2 was detected with the Chinese Center for Disease Control and Prevention (CDC) recommended kit (BioGerm, Shanghai, China), following WHO guidelines for qRT-PCR.^17^ The primers used have been described previously.^12^

### Definitions

According to the New Coronavirus Pneumonia Prevention and Control program (6th edition),^16^ COVID-19 is classified into four types including mild, moderate, severe, and critical pneumonia. Mild pneumonia means asymptomatic infection or mild clinical symptoms without abnormal chest imaging findings. Moderate pneumonia is defined as having both clinical symptoms and abnormal chest imaging findings. Patients are diagnosed as severe pneumonia when disease progresses to meet any of the following conditions: (1) significantly increased respiration rate: RR□≥□30/min; (2) oxygen saturation□≤□93% in the rest state; (3) PaO2/FiO2 ≤ 300mmHg (1mmHg = 0.133kPa). Critical pneumonia occurs when disease progresses rapidly with any of the following conditions: (1) respiratory failure which requires mechanical ventilation; (2) shock; (3) other organ failures in need of intensive care unit (ICU) monitoring and treatment. Fever was defined when the axillary temperature was equal to or above 37.5°C. Hospital stay was described as the length of stay of patients at the hospital. Neutrophil-to-lymphocyte ratio (NLR) was measured by the absolute neutrophils count to lymphocytes count. Lymphocyte-to-monocyte ratio (LMR) was measured by absolute lymphocytes count to monocytes count. Platelet-to-lymphocyte ratio (PLR) was calculated by the platelets count to lymphocytes count. Systematic immune-inflammation-based prognostic index (SII) was calculated by platelets/lymphocytes × neutrophils. Aspartate aminotransferase-to-platelet ratio index (APRI) was calculated by aspartate aminotransferase (AST) /upper limit of normal range/platelets (10^9^/L) × 100 (the upper limit of normal for AST was 40 U/L). Aminotransferase-to-neutrophil ratio index (ANRI) was measured by AST/absolute neutrophils count.

### Statistical analysis

Continuous variables with normal distribution were expressed as mean ± standard deviation (SD) and compared with analysis of variance (ANOVA). Continuous data with skewed distribution were presented as median (interquartile range, IQR) and compared with Wilcoxon rank sum test. Categorical variables were summarized as counts (percentages) and compared using the chi-square test or Fisher’s exact test. Missing data was not imputed, and was indicated in tables. Subgroup analysis was conducted by gestational period. All the analyses were performed with SPSS software, version 23. P value less than 0.05 was considered statistically significant.

## Results

### Clinical characteristics in pregnant and non-pregnant women with COVID-19

A total of 66 women of childbearing age (aged 20-40) with COVID-19 were included in our study. The mean age was 31·0±4.3 years in the pregnant group (n = 31) and 32·9±4·8 in the non-pregnant group (n = 35). Among them, 14 (21·2%) patients were aged 35-40 years (advanced maternal age). Five (14·3%) non-pregnant patients were previously infected with hepatitis B virus, and 3 out of 31 pregnant patients reported a medical history of gestational hypertension, diabetes and cardiovascular disease, separately (Table S1). There was no history of other diseases in these patients.

The clinical symptoms of the two groups were compared in Table 1. Fever was the most common symptom in both groups, but pregnant patients had a significantly lower proportion of fever than non-pregnant patients (54·8% vs. 87·5%, *p*=0·006). Moreover, pregnant patients had a shorter average interval from onset to hospitalization than non-pregnant patients (7·80 ±7·0d vs. 13·2 ± 8·2d, *p*=0·005), while a higher proportion of pregnant patients was diagnosed as severe or critical COVID-19 than non-pregnant patients (32·3% vs. 11·4%, *p*=0·039). At present, 52 (78·8%) patients were discharged out of hospital, including 22 (71%) pregnant patients and 30 (85.7%) non-pregnant patients. Mean hospital stay, heart rate, and respiratory rate were not statistically different between these two groups. Mean arterial pressure was lower in pregnant patients than non-pregnant patients (87·2 ± 10·4 vs. 96·7 ± 7·6 mmHg, *p*< 0·001).

**Table 1.**
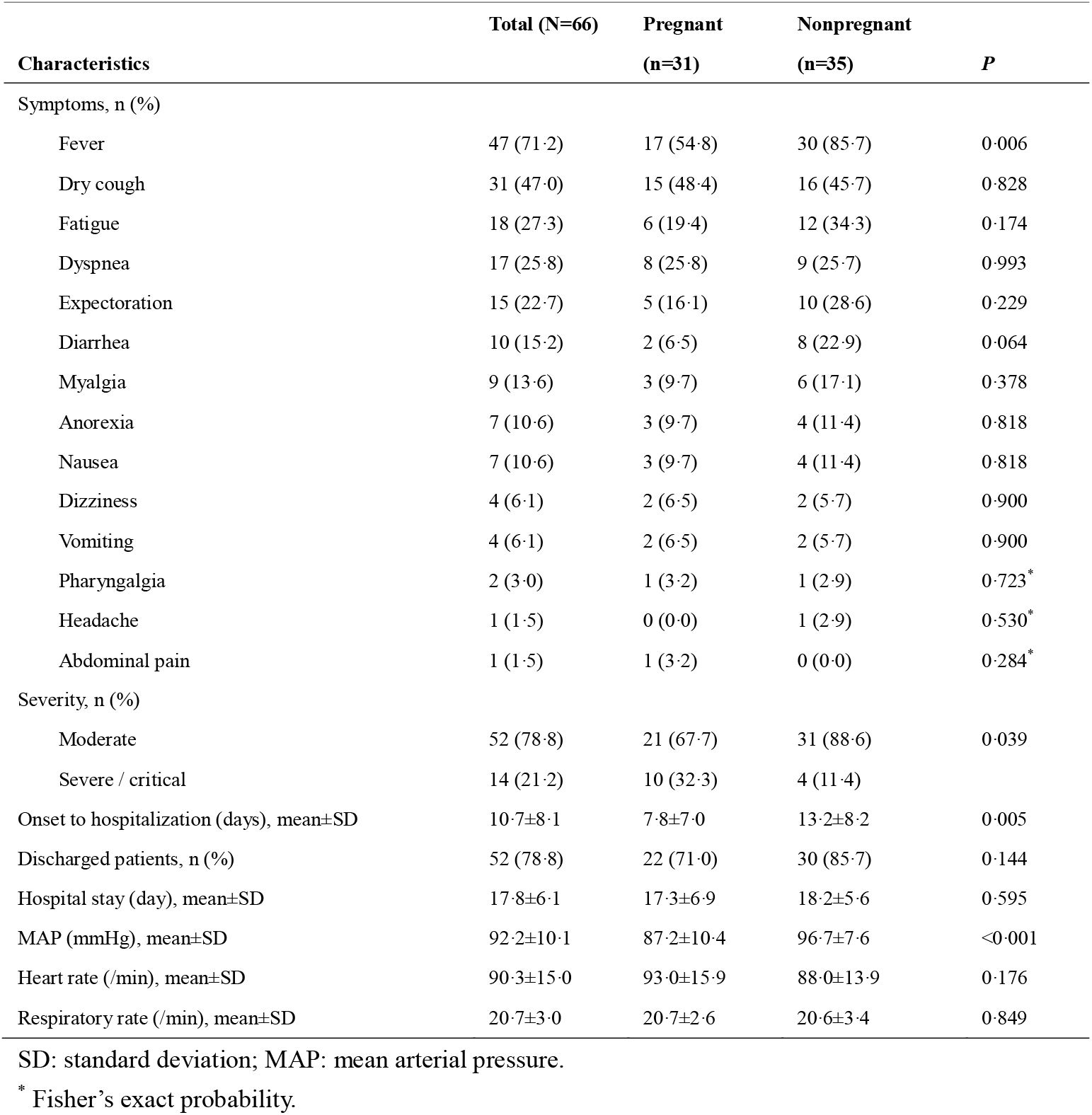
Clinical characteristics of pregnant vs. nonpregnant women of childbearing age infected with COVID-19.

Laboratory data demonstrated an increase of white blood cell (WBC) and neutrophil counts in pregnant than non-pregnant patients on admission (WBC count [10^9^/L], 6·95 ± 2·98 vs. 5·11 ± 2·13, *p*=0·005; neutrophil count [10^9^/L], 5·82 ± 2·59 vs. 3·21 ± 1·89, *p*< 0·001) (Table 2). Prothrombin time and activated partial thromboplastin time on admission were shorter in pregnant than non-pregnant patients (prothrombin time [s], 11·9 ± 1·5 vs. 13·6 ± 0·7, *p*< 0·001; activated partial thromboplastin time [s], 36·1 ± 4·1 vs. 39·6 ± 5·1, *p*=0·005), while D-dimer level were higher in pregnant than non-pregnant patients (0·91, IQR [0·58-1·32] vs. 0·36, IQR [0·26-0·71], *p*=0·003). In addition, pregnant patients were more likely to have a higher level of procalcitonin on admission than non-pregnant patients (30 [96·8%] vs. 21 [65·6%], *p*=0·002). Level of aspartate aminotransferase were increased in nine (13%) patients, including seven (22·6%) pregnant and two (5·7%) non-pregnant patients (Table 2). Though the concentrations of alanine transaminase and serum creatinine were statistically significant, there was no difference in the proportion of abnormal levels of alanine transaminase and serum creatinine between these two groups. All the enrolled patients underwent the chest CT scan (Fig. S1). Sixty-four (97%) patients showed patchy shadows or ground glass, consisting of 30 (96·8%) pregnant and 34 (97·1%) non-pregnant patients. In addition, pregnant and non-pregnant women were administered antiviral therapy (32·2% vs. 57·1%), glucocorticoid therapy (35·4% vs. 14·3%) and oxygen therapy (64·5% vs. 42·8%).

**Table 2.** Laboratory and radiological findings in pregnant vs. nonpregnant women of childbearing age infected with COVID-19.

| Laboratory indicators | Total (N=66) | Pregnant (n=31) | Nonpregnant (n=35) | P |
| --- | --- | --- | --- | --- |
| WBC ($\times 10^9/L$), mean ± SD | 5.97±2.70 | 6.95±2.98 | 5.11±2.13 | 0.005 |
| < $4 \times 10^9/L$, n (%) | 17 (25.8) | 4 (12.9) | 13 (37.1) | 0.027 |
| 4–$10 \times 10^9/L$, n (%) | 43 (65.2) | 22 (71.0) | 21 (60.9) | |
| > $10 \times 10^9/L$, n (%) | 6 (9.1) | 5 (16.1) | 1 (2.9) | |
| Neutrophil count ($\times 10^9/L$), mean±SD | 4.19±2.46 | 5.28±2.59 | 3.21±1.89 | <0.001 |
| < $1.8 \times 10^9/L$, n (%) | 7 (10.6) | 0 (0) | 7 (20.0) | 0.017 |
| 1.8–$6.3 \times 10^9/L$, n (%) | 47 (71.2) | 21 (67.7) | 26 (74.3) | |
| > $6.3 \times 10^9/L$, n (%) | 12 (18.2) | 10 (32.3) | 2 (5.7) | |
| Lymphocyte count ($\times 10^9/L$), mean±SD | 1.32±0.54 | 1.24±0.57 | 1.40±0.50 | 0.243 |
| Monocyte count, mean±SD | 0.38±0.16 | 0.36±0.18 | 0.40±0.14 | 0.331 |
| Platelet count ($\times 10^9/L$), mean±SD | 218.9±66.6 | 206.0±57.8 | 230.3±72.4 | 0.141 |
| PT (sec), mean±SD <sup>#</sup> | 12.8±1.4 | 11.9±1.5 | 13.6±0.7 | <0.001 |
| <11 sec, n (%) | 1 (1.6) | 1 (3.3) | 0 (0.0) | <0.001 |
| 11–13 sec, n (%) | 32 (50.0) | 26 (86.7) | 6 (17.6) |  |
| >13 sec, n (%) | 31 (48.4) | 3 (10.0) | 28 (82.4) |  |
| APTT (sec), mean±SD <sup>#</sup> | 37.9±4.9 | 36.1±4.1 | 39.6±5.1 | 0.005 |
| D-dimer (mg/L), median (IQR) <sup>#</sup> | 0.60 (0.31–1.02) | 0.91 (0.58–1.32) | 0.36 (0.26–0.71) | 0.003* |
| > 0.5 mg/L, n (%) <sup>#</sup> | 31 (47.7) | 25 (80.6) | 12 (35.3) | <0.001 |
| Procalcitonin (ng/ml), median (IQR) <sup>#</sup> | 0.05 (0.03–0.08) | 0.08 (0.05–0.32) | 0.04 (0.02–0.04) | <0.001* |
| > 0.1 ng/ml, n (%) <sup>#</sup> | 51 (81.0) | 30 (96.8) | 21 (65.6) | 0.002 |
| AST (U/L), median (IQR) | 19.0 (16.0–31.0) | 23.0 (18.0–40.0) | 18.0 (15.0–22.0) | 0.010* |
| > 40 U/L, n (%) | 9 (13.6) | 7 (22.6) | 2 (5.7) | 0.046 |
| ALT (U/L), median (IQR) | 16.0 (10.0–31.0) | 24.0 (10.0–35.0) | 11.0 (8.0–24.0) | 0.032* |
| > 40 U/L, n (%) | 10 (15.2) | 6 (19.4) | 4 (11.4) | 0.370 |
| Serum creatinine ($\mu\text{mol/L}$), mean±SD | 49.9±8.5 | 46.5±7.0 | 52.9±8.7 | 0.002 |
| BUN (mmol/L), mean±SD | 2.91±0.90 | 2.92±0.96 | 2.89±0.87 | 0.896 |
| LDH (U/L), mean±SD | 208.0±58.2 | 199.7±54.7 | 215.3±61.0 | 0.280 |
| CRP (mg/L), median (IQR) <sup>#</sup> | 3.9 (1.1–23.1) | 6.6 (1.4–32.2) | 3.4 (0.5–11.6) | 0.338* |
| IL-6 (pg/ml), median (IQR) <sup>#</sup> | 2.90 (1.06–4.75) | 3.7 (2.6–5.6) | 1.1 (1.1–3.1) | <0.001* |
| NLR, median (IQR) | 2.58 (1.85–4.73) | 4.21 (2.88–5.61) | 2.31 (1.33–2.54) | <0.001* |
| LMR, median (IQR) | 3.58 (2.75–4.71) | 3.55 (2.74–5.00) | 3.59 (2.76–4.37) | 0.787* |
| PLR, median (IQR) | 160.3 (135.6–226.4) | 156.8 (144.4–246.2) | 170.9 (122.0–211.0) | 0.612* |
| SII, median (IQR) | 567.6 (376.2–968.0) | 841.1 (543.2–1175.4) | 464.9 (235.7–761.0) | 0.001* |
| ANRI, median (IQR) | 6.25 (3.53–10.29) | 4.30 (3.01–10.03) | 6.96 (4.83–10.47) | 0.110* |
| APRI, median (IQR) | 0.24 (0.18–0.37) | 0.33 (0.24–0.42) | 0.21 (0.16–0.29) | 0.001* |
| Distribution of patchy shadows<br>or ground glass opacity, n (%) | 64 (97.0) | 30 (96.8) | 34 (97.1) | 0.527 |

SD: standard deviation; IQR: interquartile range; WBC: white blood cell; PT: prothrombin time; APTT: activated partial thromboplastin time; ALT: Alanine transaminase; AST: aspartate aminotransferase; BUN: blood urea nitrogen; LDH: lactic dehydrogenase; CRP: C-reactive protein; IL: interleukin; NLR: neutrophil-to-lymphocyte ratio; LMR: lymphocyte-to-monocyte ratio; PLR: platelet-to-lymphocyte ratio; SII: systemic Immune-inflammation Index; ANRI: AST-to-neutrophil ratio index; APRI: AST-to-platelet ratio index.
\* Tested by Wilcoxon rank sum test.
<sup>#</sup> Missing data: PT (n=2), APTT (n=3), D-dimer (n=1), procalcitonin (n=3), CRP (n=1), IL-6 (n=3).

### Inflammatory indices between moderate and severe/critical of pregnant and non-pregnant patients with COVID-19

Considering that inflammatory cytokine storm was the main lethal factor of infectious pneumonia such as SARS and Middle East Respiratory Syndrome (MERS),^18-21^ we compared the levels of interleukin (IL)-6 and some inflammatory indices including NLR, LMR, PLR, SII, ANRI and APRI, in pregnant and non-pregnant patients (Table 2). Higher levels of IL-6 (3·7, IQR [2·6–5·6] vs. 1·1, IQR [1·1–3·1], *p* < 0·001), NLR (4·21, IQR [2·88–5·61] vs. 2·31, IQR [1·33–2·54], *p* < 0·001), SII (841·1, IQR [53·2–1175·4] vs. 464·9, IQR [235·7–761·0], *p* < 0·001) and APRI (0·33, IQR [0·24–0·42] vs. 0·21, IQR [0·16–0·29], *p*=0·001) were found in pregnant patients compared with non-pregnant patients. Evaluated IL-6 has been reported to be positively associated with the severity of COVID-19.^22,23^ We reasoned that increased levels of NLR, SII and APRI maybe played the same roles as IL-6. Through the analysis, we found that only NLR and SII, but not other inflammatory indices, such as IL-6 and APRI, were significantly higher on admission in severe/critical pneumonia group than moderate pneumonia group (Fig. 1A-D). These results suggested that NLR and SII played a critical role in evaluating the severity of COVID-19.

**Figure 1.**
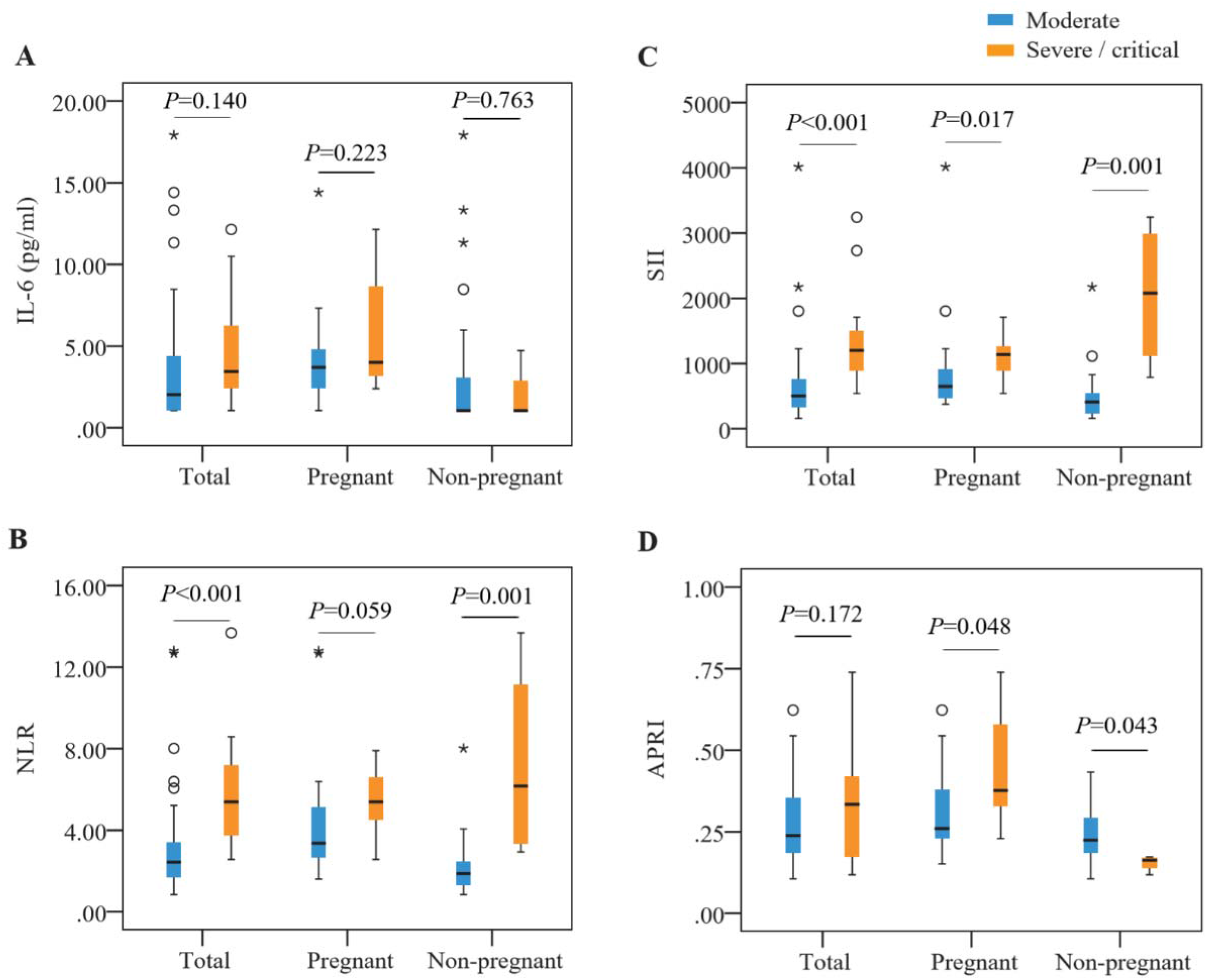
Correlation between inflammatory indices and severity of COVID-19. Interleukin (IL)-6 (**A**), systematic immune-inflammation-based prognostic index (SII) (**B**), neutrophil-to-lymphocyte ratio (NLR) (**C**), aspartate aminotransferase-to-platelet ratio index (APRI) (**D**) levels in general population, pregnant and non-pregnant women between moderate and severe/critical COVID-19.

### Characteristics of pregnant women with COVID-19 and newborns

Of these 31 pregnant patients with COVID-19, five (16·1%) were asymptomatic and one (3·2%) had an initial symptom of diarrhea. All the patients underwent fetal ultrasound examinations and no obvious abnormalities were found. These 31 pregnant patients were in the different trimesters, four (12·9%) in first trimester, five (16·1%) in second trimester and 22 (71%) in third trimester (Table 3). Among the four patients in first trimester, three underwent uterine curettage by their own choice and one voluntarily chose fetal monitoring which was also chosen by five patients in second trimester and five patients in third trimester. Among the other third trimester patients, 13 chose cesarean section voluntarily, and four gave vaginal delivery. Finally, 17 livebirths were recorded. We collected breastmilk (n=14), amniotic fluid (n=2), placenta (n=2), neonatal throat swab (n=17) and neonatal anal swab (n=5) to test the presence of SARS-CoV-2 by RT-PCR. SARS-CoV-2 was not detected in all these samples.

**Table 3.**
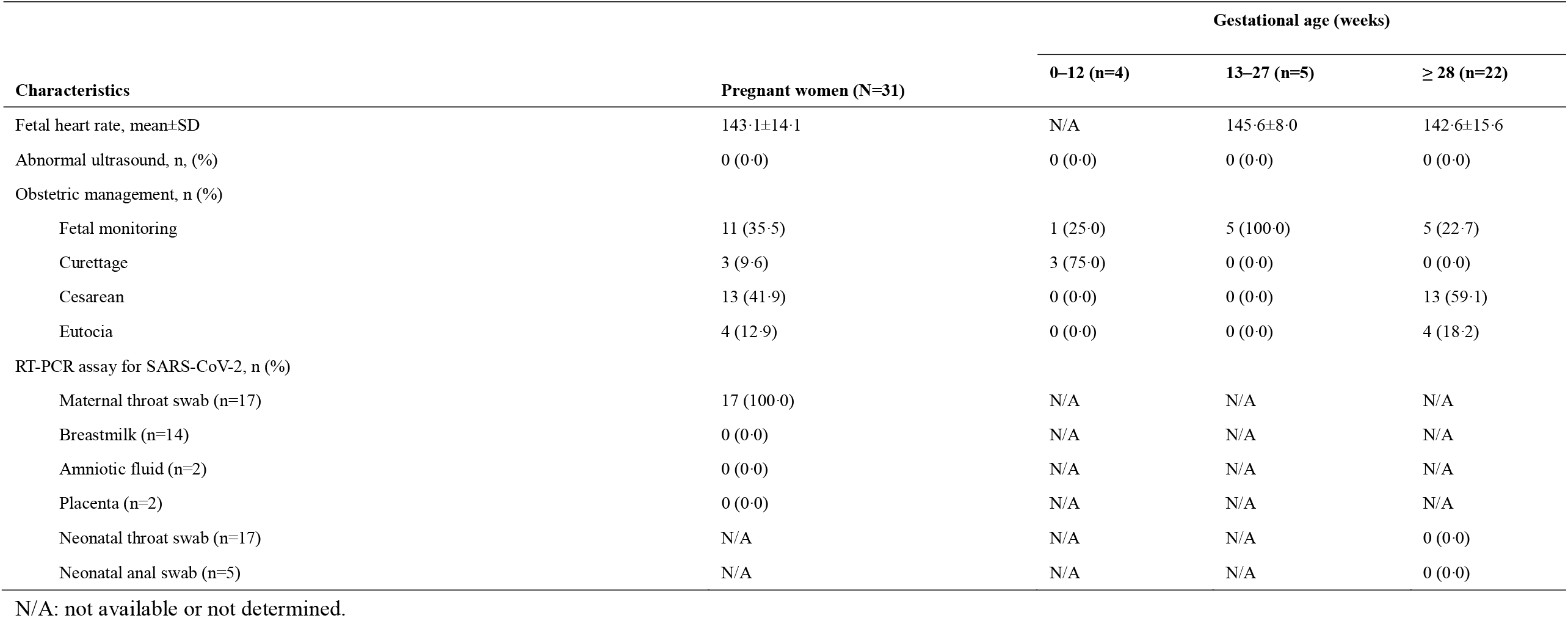
Comparison of characteristics of pregnant women with COVID-19 and newborn.

| Characteristics | Pregnant women (N=31) | Gestational age (weeks) |  |  |
| --- | --- | --- | --- | --- |
|  |  | 0–12 (n=4) | 13–27 (n=5) | ≥ 28 (n=22) |
| Fetal heart rate, mean±SD | 143.1±14.1 | N/A | 145.6±8.0 | 142.6±15.6 |
| Abnormal ultrasound, n, (%) | 0 (0.0) | 0 (0.0) | 0 (0.0) | 0 (0.0) |
| Obstetric management, n (%) |  |  |  |  |
| Fetal monitoring | 11 (35.5) | 1 (25.0) | 5 (100.0) | 5 (22.7) |
| Curettage | 3 (9.6) | 3 (75.0) | 0 (0.0) | 0 (0.0) |
| Cesarean | 13 (41.9) | 0 (0.0) | 0 (0.0) | 13 (59.1) |
| Eutocia | 4 (12.9) | 0 (0.0) | 0 (0.0) | 4 (18.2) |
| RT-PCR assay for SARS-CoV-2, n (%) |  |  |  |  |
| Maternal throat swab (n=17) | 17 (100.0) | N/A | N/A | N/A |
| Breastmilk (n=14) | 0 (0.0) | N/A | N/A | N/A |
| Amniotic fluid (n=2) | 0 (0.0) | N/A | N/A | N/A |
| Placenta (n=2) | 0 (0.0) | N/A | N/A | N/A |
| Neonatal throat swab (n=17) | N/A | N/A | N/A | 0 (0.0) |
| Neonatal anal swab (n=5) | N/A | N/A | N/A | 0 (0.0) |

N/A: not available or not determined.

Additionally, five of these 17 neonates were premature but all were beyond 35 gestational weeks (Table 4). One neonate at 35 gestational weeks had a low birthweight of 2450g, with a 1-min Apgar score of seven and a 5-min Apgar score of eight. Other neonates had a one-min Apgar score of eight and a 5-min Apgar score of nine. All these newborns were in a stable condition. No fetal death, neonatal death or neonatal asphyxia were observed.

**Table 4.** Neonatal outcome.

| Characteristics | Parturient puerpera (N=17) |  |
| --- | --- | --- |
|  | Premature delivery (n=5) | Full-term birth (n=12) |
| Gestational age at delivery (weeks), median (range) | 37 (35-37) | 39 (38-41) |
| Birthweight (g), median (range) | 2580 (2450-3130) | 3035 (2830-4100) |
| Low birthweight, n (%) (<2500g) | 1 (20.0) | 0 (0.0) |
| Apgar score |  |  |
| 1 min, median (range) | 8 (7-8) | 8 |
| 5 min, median (range) | 9 (8-9) | 9 |
| Severe neonatal asphyxia | 0 (0.0) | 0 (0.0) |
| Neonatal death | 0 (0.0) | 0 (0.0) |
| Fetal death or stillbirth | 0 (0.0) | 0 (0.0) |

## Discussion

Coronaviruses such as SARS-CoV and MERS-CoV, can cause severe adverse pregnancy outcomes, including miscarriage, intrauterine growth restriction, premature delivery, neonatal asphyxia and maternal death.^15,24-26^ However, the effects of SARS-CoV-2 infection on pregnancy is unclear, and so far there is no evidence of severity for mothers and newborns. Therefore, a cohort study is necessary and urgent to provide a reliable guideline for management of pregnant women with SARS-CoV-2 infection. To this end, we reported a cohort analysis of clinical outcomes in 31 pregnant women and 35 non-pregnant women with laboratory confirmed COVID-19. To our knowledge, this is the first cohort study that has been reported to date. Previously, a descriptive study demonstrated that the clinical characteristics of pregnant patients with COVID-19 resembled those of non-pregnant patients with COVID-19.^12^ However, we found a shorter interval from onset to hospitalization and severer COVID-19 in pregnant patients than non-pregnant patients with SARS-CoV-2 infection. This phenomenon could be partly explained by the special states of immune suppression and physiological adaptive changes during pregnancy. Infectious pneumonia is an essential cause of morbidity and mortality in pregnant women. For example, H1N1 virus caused a higher rate of hospital admission and an increasing risk for pregnancy complications in pregnant women than the general population.^27^ In our study, despite no death in pregnant women and neonates, our finding suggested that more attention should be paid to the pregnant women especially to those who had a history of exposure to the confirmed cases.

Additionally, pregnant women had insidious and atypical symptoms which might increase the risk of misdiagnosis. Fever was the most common symptom in the general population, as previously reported (over 80%).^28,29^ In our study, we found the similar pattern of symptoms in pregnant women, while the proportion of fever in pregnant women was far lower than that in non-pregnant women (54·8% vs. 87·5%, *p* = 0·006). Temperature screening has been the most proposed and useful method to identify the suspected patients in population. Thus, a low proportion of fever might increase the risk of missing the suspected patients in pregnant women. Furthermore, five (16·1%) pregnant women who were diagnosed by laboratory confirmation of SARS-CoV-2 did not have any initial symptoms but had the history of exposure, and one (3·2%) pregnant woman had an initial symptom of diarrhea. Importantly, chest CT scan of all patients showed moderate to severe inflammation in lungs. These findings plainly indicated that it was not enough to screen pregnant women for COVID-19 based solely on clinical symptoms. Monitoring, early detection and diagnosis are urgent and necessary for pregnant women, especially for those with a history of exposure.

There were numerous differences in the laboratory findings between pregnant patients and non-pregnant patients. Compared with nonpregnant patients, pregnant patients had more white blood cell and neutrophil counts, shorter prothrombin time and activated partial thromboplastin time, higher level of D-dimer, procalcitonin and aspartate aminotransferase. Moreover, there was no statistical difference in the proportion of abnormal levels of alanine transaminase and serum creatinine between these two groups, while pregnant patients had higher level of alanine transaminase and lower level of serum creatinine than non-pregnant patients. These abnormalities suggested that pregnant patients might have diminished cell-mediated immunity, hypercoagulable state, and increased hepatic injury compared with non-pregnant patients. These abnormalities were not unique between pregnant and non-pregnant patients with COVID-19. Previous studies showed that pregnant patients were more likely to have complications and adverse outcomes such as sepsis and hepatic injury with SARS-CoV and MERS-CoV infection.^30,31^

Monitoring the severity of COVID-19 is imperative for the adjustment of treatment strategies. Though analyzing cytokines in peripheral blood of 123 hospitalized patients with COVID-19, Wan et al. found that IL-6 were higher in severe patients than in mild patients.^23^ Another team observed the similar phenomenon and further demonstrated significant decrease of IL-6 after treatment for patients with severe COVID-19.^22^ Our study showed that pregnant patients had increased IL-6 level compared with non-pregnant patients, which is consistent with our previous finding that more pregnant patients were diagnosed as severe or critical pneumonia than non-pregnant patients. However, in our study, although the median level of IL-6 was higher in severe/critical group than moderate group, no significant difference was identified between these two groups. NLR and SII are the inflammatory markers which have been reported to be associated with the progress of disease.^32,33^ We found that NLR and SII were positively correlated with the severity of COVID-19. It seems that these two inflammatory indices have a higher sensitivity than IL-6 on evaluating the severity of COVID-19. Besides, unlike IL-6, NLR and SII could be easily calculated from blood standard examination. A large, prospective study is necessary to investigate the value of NLR and SII in monitoring the severity of COVID-19.

Another focus of this study was to evaluate the possibility of vertical transmission of COVID-19. We did not find evidence of SARS-CoV-2 by RT-PCR in breastmilk, amniotic fluid, placenta, neonatal throat swab, and neonatal anal swab. These findings suggested that SARS-CoV-2 could not penetrate the placental and blood-milk barriers. This observation confirms 5 premature delivery and 12 full-term birth including 17 pregnant patients and 17 neonates, showing that COVID-19 was negative in all the neonates. Moreover, we reviewed the previous reports about pregnant women with SARS-CoV and MERS-CoV infection. There was no evidence that SARS-CoV and MERS-CoV could be transmitted from infected mothers to infants during pregnancy.

This study has several limitations. First, we only included the hospitalized patients with laboratory confirmed COVID-19. It could be more methodologically sound to include both pregnant outpatients and inpatients in the analysis to obtain a lees biased estimate comprehensive understanding of the effect of COVID-19 on pregnancy. Second, patient’s information such as pregnant outcomes in first and second trimester were incomplete and unavailable at the time of analysis. Third, selection bias might be introduced using 35 non-pregnant patients of reproductive age as control cohort. To clarify, we made a baseline comparison between these 35 non-pregnant patients and all female patients of reproductive age from Hunan province near Wuhan where COVID-19 broke out and found no difference (Table S2). Finally, considering the limited number in this study, we should treat the results with more caution. Non-significant P value still could not rule out the possibility of false negative results when comparing pregnant and non-pregnant patients.

In summary, pregnant women with COVID-19 had insidious and atypical symptoms which increased the risk of misdiagnosis. SII and NLR could be useful markers to evaluate the severity of the COVID-19. There was no evidence of vertical transmission during pregnancy. These findings will be crucial to inform prophylactic and therapeutic strategies for pregnant women with COVID-19.

## Data Availability

All data referred to in the manuscript,

## Contributors

MY and XC designed the study. LZ, CF, SY and HC collected the data. MY and MS were involved in data cleaning, and verification. MY, MS, and GD analyzed the data. GD and MY drafted the manuscript. MY, GD and MS contributed to the interpretation of the results and critical revision of the manuscript for important intellectual content and approved the final version of the manuscript. All authors have read and approved the final manuscript. MY, XC SY and HC are the study guarantors. DB independently reviewed all data and analysis due to my expertise in emerging infectious disease.

## Data sharing

No additional data are available.

## Acknowledgements

We thank all the hospital staff members for their efforts in collecting the information that used in this study; thank the patients who participated in this study, their families, and the medical, nursing, and research staff at the study centers.

